# Risk tolerance and risk perception as determinants of successful and sustainable hospital admission avoidance interventions: a realist review

**DOI:** 10.1101/2025.07.14.25331501

**Authors:** Claire Maynard, Alyson Huntley, Helen Baxter, Matthew Booker

**Author notes:** Corresponding author: Matthew Booker.

## Abstract

**Background:** Health systems under pressure from rising demand for elective and emergency care are prioritising the reduction of avoidable hospital admissions and the expansion of urgent community care. Yet, inconsistent service provision and fragmented, short-term initiatives have resulted in variable patient experiences, inequities, and inefficient use of resources. The mechanisms driving the success or failure of hospital admission avoidance interventions (HAAIs) remain unclear. This study aimed to develop initial programme theories to understand what makes HAAIs effective, for whom, and in what contexts.

**Methods:** A realist review was undertaken to generate explanatory insights into how complex interventions function in practice. Peer-reviewed and grey literature were eligible for inclusion, with a focus on recent publications. A reverse chronological quota sampling approach was used for screening, supplemented by citation tracking and stakeholder recommendations. Empirical evidence, middle-range theories, and public and professional stakeholder input were synthesised to iteratively develop and refine initial programme theories.

**Results:** From 3,395 records, 68 studies were included in the synthesis. Risk emerged as a central concept shaping how HAAIs are experienced by patients, carers, and healthcare professionals.. Three initial programme theories were developed, each exploring how risk can be managed and navigated in HAAIs: (1) promoting patient and carer acceptance of alternatives to hospital admission (2) nnavigating clinical complexity and uncertainty, and (3) clarifying clinical and organisational roles and responsibilities. Contextual factors, including individual attitudes, interpersonal trust, and organisational risk appetite, were found to influence how these mechanisms operate.

**Conclusion:** HAAIs may be most effective when tailored to local contexts, balancing hierarchical and relational mechanisms to manage and navigate risk for patients, carers and healthcare professionals.

## Background

Unscheduled hospital admissions can have negative consequences for patients and carers, while placing significant strain on health systems. Reducing these admissions offers an opportunity to improve patient outcomes and experiences, and to alleviate pressure on secondary care services (1). This priority has gained further urgency in the aftermath of the COVID-19 pandemic, which raised public awareness of the risks associated with hospital stays (2). In response, health systems are increasingly focusing on managing urgent care-sensitive conditions (UCSCs) outside hospital settings, shifting care towards community and home-based environments. However, enabling this shift requires fundamental changes in how complex, non-emergency conditions are identified and managed (3).

Hospital admission avoidance interventions (HAAIs) span a diverse range of services and models, from enhanced emergency department triage to community-based step-up care and hospital-at-home schemes. These interventions vary widely in design, implementation, and maturity across local health systems (8, 9). While some evidence supports their effectiveness for specific patient groups, many HAAIs struggle to demonstrate sustained impact or scalability. Traditional study designs have offered limited insight into why some interventions succeed while others do not (4–6)(7).

To inform the design, implementation, and scaling of effective HAAIs, a deeper understanding is needed of how they work, for whom, and in what contexts. This study used a realist review approach to examine existing evidence and develop initial programme theories (IPTs). Realist approaches are increasingly used in health services research to unpack how complex interventions interact with local contexts to produce outcomes (10)(11). Unlike traditional effectiveness studies, realist methods seek to explain the underlying mechanisms at play, offering insights into causality and context-specific success factors (12).

## Methods

This realist review was written to RAMESES publication standards (13) and the protocol was prospectively registered on PROSPERO (ID: CRD42023468852).

### Inclusion and exclusion criteria

An initial purposive scoping review of national and regional policy documents on urgent and emergency care in England, along with discussions with an external realist review expert (JJ), informed the development of a spatio-temporal framework of hospital admission avoidance from a system perspective: 1) awareness of candidacy for hospital admission avoidance; 2) access to alternative services; 3) clinical decision-making for hospital admission avoidance (HAA); and 4) provision of urgent care in out-of-hospital settings. We used this framework to guide the development of eligibility criteria for empirical literature and a database search strategy (see Supplementary material).

The term *urgent care-sensitive conditions* (UCSCs) was used to define the population, or condition, of interest, where UCSCS are acute exacerbations or urgent conditions that could be managed without hospital admission (14) (1). This term acknowledges the contextual nature of avoidable hospital admissions, which can depend on local service availability, patient condition, social environment, and patient or carer preferences. Studies exploring the outcomes or experiences of patients or informal carers with UCSCs as well as health and care professionals (HCPs) involved in any stage of care or implementation of a HAAI were eligible populations. Case management interventions for people with chronic conditions were excluded. Consistent with the realist approach, outcomes were not specified in advance. All study types were eligible for inclusion, with a focus on explanatory qualitative evidence. A publication date limit of 1 January 2019 was set as the research team determined that the COVID-19 pandemic had significantly impacted the policy landscape and broader context surrounding admission avoidance, making more recent literature a priority.

### Searching and screening

Four main search techniques were used to collect empirical evidence: a structured database search; websites of relevant organisations for grey-literature (The King’s Fund, The Health Foundation, Nuffield Trust, Healthwatch); citation pearling; and manual identification of studies known to members of the research team and stakeholders. The following databases were searched on 16^th^ October 2023 and updated on 2^nd^ April 2024 using a search strategy developed with a subject expert librarian (supplementary material): MEDLINE, CINAHL, PsycINFO, Embase, Cochrane Library (CCRCT & CDSR), Scopus, Web of Science and the pre-print servers, medRxiv. For the database search results, reverse chronology quota screening was used to prioritise the most recent information (Stott 2024; Taylor 2024).

One reviewer screened articles by title and abstract, starting with the most recent publications, until approximately 15 articles were selected for full text review. Two reviewers then independently screened the full texts, with any disagreements resolved with a third reviewer. This was repeated three times, resulting in approximately 60 articles being included. Covidence, an online reviewing platform, was used to screen the articles retrieved from database searches. Following realist quality assessment principles, selected full-text articles were appraised during synthesis based on their relevance and depth of insight to the developing programme theory (12). Eight articles of low relevance were excluded, including studies on psychiatric urgent care (due to differing policy context governing clinical decision-making for hospital admission avoidance); interventions for detecting exacerbations of chronic conditions (as these populations did not have an ‘active’ UCSC), and HAAIs designed for people with COVID-19. Exclusion decisions were discussed within the research team.

### Realist synthesis and theory development

The technique of appraisal journalling guided data extraction and analysis (15). The lead reviewer (CM) documented reflections on the article’s findings in relation to the initial framework, while further insights were gained through team discussions, drawing on HAAI expertise. Descriptive data on intervention components, implementation approaches, and measured outcomes were extracted. A batch approach to article selection and analysis enabled iterative development of IPTs, allowing reviewers to assess whether subsequent articles supported, challenged, or added to emerging findings. The realist context-mechanism-outcome (CMO) heuristic guided the development of causal relationships, with mechanisms conceptualised as resource-response interactions – intervention components as resources and stakeholders’ behavioural or cognitive processes and actions as responses (16).

Using a retroductive approach (27), emerging IPTs were informed and structured using existing middle-range theory and mechanisms identified in the empirical and theoretical literature. Analysis also considered how individual, interpersonal, and institutional contexts shaped these interactions and outcomes.

### Stakeholder involvement and theory refinement

Stakeholder engagement supported the refinement of programme theory throughout the review. Six public contributors with lived experience of hospital admission avoidance, as patients and/or informal carers, were consulted during the scoping phase to inform the IPTs and later during discussions of emerging findings.

Semi-structured realist interviews were conducted with six professional stakeholders, purposively selected for their experience in the design and/or implementation of HAAIs. Participants were recruited via personal networks and snowball sampling. Professional stakeholder roles included: Medical Director of a Hospital at Home Service; ICB Director (Nurse); Acute Hospital Physician; Community Interface Lead GP; NHS England Programme Director; Deputy Director of Social Care (regional). Verbal informed consent was obtained from each participant and ethical approval was obtained from the University of Bristol faculty ethics committee.

## Results

### Summary of studies

A total of 68 articles were included in the review: 53 from database searches (including 11 conference abstracts), eight articles from citation pearling or manual selection, and seven grey literature articles (see supplementary material for a study selection diagram). To map the wide diversity and range of HAAIs, we categorised them according to our initial process framework, however interventions primarily fell into either (1) supporting clinical decision-making for HAA and (2) provision of urgent care in out-of-hospital settings (see Table 1). No interventions were identified solely addressing awareness of candidacy or access to alternative services. Eleven sub-categories were created based on more defined intervention strategies, however, due to the multi-faceted nature of interventions, some studies appear in more than one sub-category.

**Table 1.** HAAI interventions and intervention strategies.

| Hospital admission avoidance interventions | References |
| --- | --- |
| **Supporting clinical decision-making** |  |
| Remote urgent advice, guidance and/ or referral service for patients and/ or carers | (28, 29, 47, 54, 61, 70, 81) |
| Enhanced paramedic assessment | (37, 51, 67, 91-93) |
| Enhanced assessment (and management) of urgent care sensitive conditions in care homes | (32, 39-42, 46, 49, 55, 72, 83, 89, 90) |
| Specialist assessment for HAA in emergency departments | (34-36, 38, 53, 59, 64, 68, 69, 77, 94) |
| Emergency department dedicated personnel for HAA referrals | (31, 56, 66) |
| **Provision of urgent care in out-of-hospital settings** |  |
| Urgent care services to people's place of residence e.g., urgent community response and rapid response services | (33, 52, 57, 58, 60, 62, 63, 65, 84) |
| Urgent palliative and end-of-life care in the community | (67, 71, 95) |
| Step-up services e.g., Hospital at Home (HaH) and Virtual wards (VW) services | (7, 43, 50, 74, 75, 78, 80, 86-88) |
| Expanded primary care capacity (workforce and/or hours) | (70, 76, 85) |
| Ambulatory same day urgent care clinics e.g., Acute Respiratory Infection hub, surgical ambulatory clinic | (45, 48, 73, 82) |
| Outpatient urgent care services | (29, 30) |

The majority of studies were observational, including several service evaluations: 22 used quantitative methods (28–48), 15 qualitative (49–63) and 14 mixed methods or were quality improvement studies (64–77). Additionally, seven narrative, rapid, or scoping reviews (78–84), four qualitative or realist syntheses (7, 85–87), four systematic reviews (88–91), and two theses (one mixed-methods study and one ethnographic study) (92, 93). Among the primary qualitative and mixed-methods studies, healthcare professionals’ views were represented in 17 studies, patients’ views in 10, and carers’ views in five. Studies were conducted across 13 countries, with the majority from England (n=28).

### Use of middle-range theory

Emerging IPTs were informed and structured using existing middle-range theory and causal relationships identified in the empirical and theoretical literature. These highlighted the role of risk in the implementation, acceptance, and sustainability of new or unfamiliar services in healthcare settings, including urgent and emergency care (17–21). Relevant mechanisms included strategies to navigate uncertainty and mitigate perceived risk, enhancing risk tolerance and supporting service adoption (22–26). Drawing on sociological theory, mechanisms were further categorised as *hierarchical* (those that rely on authority, protocols, or formal accountability structures to manage risk) and *relational* (those that build trust, collaboration, and shared understanding among stakeholders to support risk navigation) (108).

The significance of risk and uncertainty was echoed by professional stakeholders, who described how unclear professional or organisational responsibilities often led to HAAIs being unsustained or failing to adapt to higher levels of clinical acuity. Several noted that differing risk appetites across provider organisations influenced how individuals and teams behaved. Public contributors described weighing the accessibility, perceived safety, and quality of alternative services against the familiarity of hospital admission, despite known risks associated with hospital stays. They also reflected on risk in terms of access and gatekeeping, highlighting a tension between individual agency - having choice and control over hospital admission avoidance options - and system-level control over who could access services.

This moved the analysis beyond the initial admission avoidance stages to identify broader explanatory themes. The following sections present three refined IPTs, illustrating how risk is managed and navigated in HAAIs, by key stakeholders: patients and carers (IPT 1), and healthcare professionals (IPT 2 and 3).

### Initial programme theories

The following section presents three refined IPTs, developed through synthesis of empirical evidence, stakeholder insights, and middle-range theory (see Box 1). Each IPT outlines the resources and strategies used to manage risk, how stakeholders respond, and the contextual factors that influence whether these mechanisms lead to effective and sustainable HAAIs across different settings.

#### Box 1

##### Summary of Initial Programme Theories (IPTs)

###### IPT 1: Promoting patient and carer acceptance of alternatives to hospital admission

– Patients and carers often perceive greater risk when accessing hospital admission avoidance (HAA) services compared to presenting to the emergency department, particularly when unfamiliar with available options. Feelings of disempowerment, confusion, and concerns about coping at home without hospital-level support can lead to delayed care and poorer outcomes. This IPT explores how interventions that promote a sense of safety and control, through accessible services, shared decision-making, home-based care, monitoring, and integrated follow-up, can reduce perceived risk in alternatives to hospital admission.

###### IPT 2: Navigating clinical complexity and uncertainty

– Hospital admission avoidance involves inherent clinical, ethical, and medico-legal risks for HCPs, particularly in urgent and emergency care settings where acuity is high and time is limited. Under such uncertainty, HCPs may default to hospital admission as the safer, more familiar option, driven by concerns about patient deterioration, professional accountability, or litigation. This IPT examines how the ability to navigate clinical complexity, supported by comprehensive assessment, senior clinical input, diagnostics, and decision-support tools, can enhance HCPs’ risk tolerance for HAA. Individual, interpersonal and institutional factors that influence whether these mechanisms are effective in different settings are also explored.

###### IPT 3: Clarifying clinical and organisational roles and responsibilities

– Managing urgent care in community and home settings often requires HCPs to work beyond their usual scope of practice and across organisational boundaries. Collaboration between services, particularly at the primary-secondary care interface and with emergency services, is crucial to avoid gaps in clinical responsibility. Stakeholders and literature highlight risks where roles are unclear, such as services holding partial responsibility without full medical oversight. This IPT explores how clearly defined roles, supported by senior oversight, clear protocols, and interdisciplinary teamwork, can enhance risk tolerance for hospital admission avoidance. It also considers the influence of inter-professional trust and organisational risk appetite on these mechanisms.

### Promoting patient and carer acceptance of alternatives to hospital admission (IPT 1)

#### Ease of access

Accessible and convenient alternatives to emergency departments (EDs) help promote patient and carer acceptance of non-hospital care by increasing their sense of control and trust in the system. Included studies and public contributors emphasised the value of visible, easy-to-navigate services— such as walk-in centres, mobile units, and self-referral options—which offer greater autonomy over when and where to seek help. These alternatives can reassure patients that timely care is available outside of hospital settings, supporting confidence in community-based options (47) (82) (45) (73) (80).

Expanding GP capacity, especially in underserved areas, alongside remote urgent care services like telephone advice and telemedicine, improves access, promotes continuity, and builds trust in non-hospital care for patients with limited mobility or living far from EDs (54, 58, 61, 76). When delivered by experienced clinicians with access to medical records, virtual consultations were associated with reduced ED use, improved patient flow, and greater satisfaction (28, 46, 47, 81, 84). For high-risk patients, rapid access to specialist advice via phone and referral to integrated same-day care pathways helped prevent unnecessary ED attendance (29, 30). However, the acceptability of remote services depends on digital access, language support, and cognitive capacity, which can present barriers for some group (54, 61).

While increased access can improve patient acceptance of non-hospital care, stakeholders and some studies highlighted concerns about unintended outcomes. Greater service visibility and convenience may risk driving up demand or duplicating existing provision, potentially straining limited resources and undermining cost-effectiveness (47). However, routine data from hospitals using different models of GPs in EDs showed increased demand over time was unrelated to service visibility (68, 69). Economic analyses have suggested that cost savings depend on referral sources, underscoring the need for careful pathway design to maximise efficiency and sustainability (48).

#### Shared-decision making

Shared decision-making can reduce perceived risk of alternatives to hospital admission for patients and carers by fostering a sense of control, reassurance, and involvement in care decisions. When supported by trusting patient-provider relationships, it creates space to openly explore preferences and address concerns about non-hospital care options, helping patients feel informed and confident in the chosen pathway (37, 49, 75, 85, 89, 95). However, inexperience or pressure may lead some staff to inappropriately shift responsibility onto patients or carers (89). While collaborative decision-making can empower patients, it may also feel burdensome, particularly in end-of-life scenarios where clearer clinical direction may be preferred (49, 60). In some cases, patients may also perceive comprehensive assessments as unnecessary or disempowering if they already have a clear care preference. However, patient and carer preferences, often informed by cultural values, can shape the perceived acceptability of out-of-hospital care. For example, in some communities, hospital care may be preferred at the end of life.

#### Home-based and person-centred care

Evidence from included studies and stakeholder contributions suggests that receiving urgent care at home or in a familiar setting can reduce perceived risks and offer several advantages over hospital admission. Patients reported feeling safer and more in control in their own environment, which supported mobility, daily routines, and reduced anxiety related to hospital-associated risks such as infection, contributing to better physical, psychological, and functional outcomes, and faster recovery (87, 88). For informal carers, avoiding the disruption of hospital stays reduced logistical burdens and helped preserve family support networks (7). However, several studies also warned that when carers’ capacity to provide care is not adequately assessed, the burden can become unsustainable, potentially leading to unplanned admissions or poorer outcomes (78, 80, 86, 87).

In home settings, several studies highlighted that HCPs are better able to address patients’ broader psychosocial needs in home settings, enabling more tailored and holistic care that could build confidence in managing conditions long-term and help prevent future admissions (7, 50, 52, 78, 86, 87) (51). Person-centred care was most effective when supported by strong, trusting relationships between patients and providers underpinned by clear communication and professional competence (96–98). This reassured patients and carers that urgent care delivered at home could be a safe and effective alternative to hospital admission (51, 87).

#### Monitoring and surveillance

Remote monitoring, such as continuous vital sign tracking or direct communication with clinicians via phone or video, can reassure patients and carers that deterioration will be detected and managed promptly, enhancing their sense of safety of home-based care (7, 58, 87). Some patients in a HaH service reported feeling more supported at home than in hospital, however, public stakeholders noted that this reassurance may not extend to those who view monitoring as intrusive or who lack the skills or confidence to use digital tools. Acceptance of these technologies depends on access to training, user-friendly design, and adequate digital infrastructure (43, 54, 78). The effectiveness of remote monitoring is also shaped by geography; people living alone or in rural areas may feel less secure, especially overnight, even with remote clinical support (44).

#### Integration and follow-up

Public and professional stakeholders, in addition to included studies, agreed that the end of an urgent care episode is often poorly defined, leaving uncertainties about follow-up care (80). Without clear handover to their regular provider, patients can feel frustrated about the fragmentation of care and unprepared to manage at home (34, 51, 86). These risks are heightened in stand-alone services lacking access to patient records or referral pathways back to primary care (Nuffield). One study found that a paramedic pathway involving GP follow-up improved care integration and reassured patients about ongoing safety (51). Senior clinical oversight to coordinate follow-up can further reduce fragmentation, support long-term condition management, and help prevent deterioration and readmission, enhancing outcomes for both patients and carers (34, 43, 58, 86).

### Navigating clinical complexity and uncertainty (IPT 2)

#### Comprehensive assessment

Comprehensive assessment pathways, such as CGA or specialist-led triage in EDs, support HCPs to navigate the clinical risk involved in hospital admission avoidance decisions. By allocating time, space, and multidisciplinary expertise, these pathways enable thorough evaluation of urgent or emergency needs, particularly in patients with complex conditions. Evidence shows they can reduce unnecessary admissions and improve patient experience (34-36, 53, 64, 66, (69). While such assessments can lengthen ED stays, this can be mitigated through dedicated spaces or streamlined processes (31, 94, 102).

However, across settings, several factors can influence the extent to which staff can carry out a comprehensive assessment. For example, paramedics and care home staff have faced challenges implementing CGA and assessing UCSCs due to insufficient training and limited time, which in some cases led to no reduction or even increases in admission rates (37, (46, 83, 49, 89). Crucial contextual enablers include staff training and education, guidance on when comprehensive assessment is required (35, 38, 94, 102) in addition to institutional changes to care pathways to allow additional time for comprehensive assessment.

#### Diagnostics and decision-support tools

Decision-support tools and point-of-care diagnostics can help HCPs manage uncertainty when assessing the need for urgent or emergency care, particularly in settings like care homes, where staff may lack clinical training or confidence (32). Evidence suggests that these tools are most effective in specific scenarios, such as ruling out serious conditions in patients without complex needs and where clinical ambiguity exists (32, 92).

However, successful implementation is highly context-dependent. Cultural resistance can arise when such tools are perceived as undermining professional judgement or failing to reflect clinical complexity (33, 69, 83, 92). Evidence suggests that their effectiveness relies on several enabling conditions, including co-development with end users, adequate training, and careful integration into established clinical workflows (7, 92). In care home settings, for example, diagnostic tools supported safe hospital admission avoidance only when accompanied by staff training and used in conjunction with clinical judgement (83). In addition, financial incentives are often needed to enable their use in community-based services and reduce reliance on emergency departments for diagnostics (41).

More broadly, the effectiveness of these tools is shaped by the attitudes and beliefs of staff. Stakeholder input and included studies emphasised that HCPs’ perceptions of clinical risk, and their confidence in the safety and appropriateness of hospital admission avoidance, vary considerably (7). These differences are influenced by professional background, local organisational culture, and wider system norms, all of which strongly affect referral behaviours. Building confidence in decision-support tools therefore requires not only technical integration, but also cultural alignment and professional trust.

#### Senior clinical oversight

Access to senior clinical or medical decision-makers plays a critical role in managing risk, especially in settings where staff lack confidence or expertise in making HAA decisions. Shifting risk responsibility to more senior staff helps prevent defaulting to emergency services as a precaution (7, 40, 86). In care homes, in-person or remote support from GPs, specialist nurses (89, 90), or consultant-led teams (40, 83) has been shown to reduce ambulance use and hospital admissions (42, 75). For paramedics, phone access to doctors during out-of-hours palliative care provided “greater confidence and certainty to treat palliative patients during out-of-hours calls” (95) and reduced hospital admissions.

However, this relies on inter-professional trust. Where hierarchies are rigid or relationships strained, staff may hesitate to hand over decisions, fearing criticism (40, 49, 83). For example, in one study, poor relationships between care home staff and acute providers led to mutual distrust, undermining the intervention (83). Such breakdowns can also result in conflicting advice, reducing patient confidence in care (87). Sustained funding and incentives are essential for recruiting and retaining physicians in community services (7, 39, 50, 89). For instance, a hospital-at-home (HaH) service was forced to lower its acuity threshold due to difficulties recruiting geriatricians (50). Flexible models, such as rotational posts, consultant hotlines, or GP input, exist but often lack long-term sustainability. Digital infrastructure and funding also underpin successful virtual urgent assessment models (41, 55, 89). For further detail on virtual assessment in clinical decision-making, see studies such as (55).

### Clarifying clinical and organisational roles and responsibilities (IPT 3)

#### Clinical guidelines and service criteria

Clear referral criteria for HAAIs (often based on risk stratification and cost-effectiveness evidence) (38, 62, 65, 75) can promote staff risk tolerance by reassuring HCPs that the level of clinical risk is appropriate and aligned with their skills and service capacity (7, 57, 65, 78, 86, 88). Professional stakeholders warned that vague or poorly defined service specifications, especially in newer models like VW and HaH, can lead to mismatches between clinical risk and available expertise, increasing safety risks. Professional stakeholders emphasise that agreeing on acceptable risk levels and clearly defining risk responsibilities across organisations are essential for effective, integrated urgent care pathways. The evidence also highlights that organisational and cultural resistance—especially in risk-averse settings—can impede the adoption of new services (7, 57, 65, 86, 89).

Formal clinical guidelines that clarify service scope or expand HCP responsibilities further support safe risk tolerance (7, 78). For example, updated guidance enabled paramedics to deliver urgent palliative care with greater confidence and fewer medico-legal concerns (95). Similarly, in nursing homes, clear protocols and training empowered staff to deliver more advanced care, reducing ED visits and hospital admissions (75).

#### Interdisciplinary teamwork and supervision

Collaborative multidisciplinary teams (MDTs) can support HCPs in sharing responsibility for clinical risk. Strong relational dynamics within teams enable professionals to navigate complexity more confidently, especially when managing high-risk cases. Informal peer support and regular MDT meetings promote knowledge exchange and collective problem-solving (50, 52, 86) (62). However, in less mature teams or those without strong leadership, weak inter-professional trust can reduce risk tolerance, leaving less experienced staff uncertain or reluctant to act (52). Stakeholders highlighted that mutual respect and role clarity are essential for teams to function effectively and safely distribute clinical risk.

Supervision from experienced colleagues can help HCPs develop the confidence and autonomy needed to manage clinical uncertainty. In urgent community response services, senior team members acted as mentors, supporting colleagues to move beyond traditional roles (52, 86). For paramedics working in general practice or delivering palliative care, GP supervision and communication with specialist teams helped shift decision-making from protocol-driven to patient-centred approaches, enabling better risk navigation (85, (95). These support structures are vital for embedding new practices and empowering staff to operate safely in complex community settings.

#### Collaborative service design and development

Co-developed service agreements that clearly define clinical, operational, and legal responsibilities help organisations navigate risk by establishing transparent roles and shared expectations (58). In virtual ward services, such agreements between secondary and community care providers reinforced a common vision and enabled integrated care delivery (86).

Similarly, in care homes, admission avoidance interventions were most successful when developed collaboratively with staff and supported by quality improvement initiatives that built inter-professional trust and buy-in (42, 55, 75, 89, 90). For example, during the early implementation of an UCR service, weekly meetings between the UCR team and the ambulance service facilitated the review and safe transfer of calls from the ambulance call stack. As inter-organisational trust grew, the acuity of referrals increased, leading to fewer ambulance dispatches and conveyances (60). Conversely, unclear or ad-hoc decision-making can foster tension and uncertainty over shifting responsibilities (7, 89). Sustainable funding and financial incentives are also essential to secure buy-in for implementation (39, 74).

## Discussion

Drawing on diverse empirical studies, stakeholder insights, and existing theory, this realist synthesis highlights risk as a core factor influencing how HAAIs work. Patients, carers, and HCPs experience various forms of risk when receiving and delivering care, respectively, and if these are not adequately addressed, hospital admission remains the default option. Descriptive decision theories, such as Ellsberg’s (101), suggest that when faced with uncertainty, individuals often opt for the most familiar or least ambiguous choice to minimise perceived risk.

### Summary of findings

This realist review identified three interrelated programme theories that highlight the conditions under which HAAIs are most likely to be effective and sustainable. Firstly, patient and carer acceptance of alternatives to hospital admission is contingent upon their perceived safety, sense of control, and trust in healthcare professionals (IPT 1). When these conditions are supported, through accessible and clearly communicated services, shared decision-making, and care delivered in familiar or home settings, patients and carers are more likely to view non-hospital options as safe and appropriate.

Secondly, the ability of HCPs to safely and confidently make decisions that avoid hospital admission is shaped by their capacity to assess and respond to clinical complexity and uncertainty (IPT 2). This requires sufficient time, appropriate diagnostic and decision-support tools, and senior clinical oversight. These resources enable staff to tolerate higher levels of clinical risk and avoid unnecessary admissions. This finding aligns with ethnographic research in ambulatory emergency care, which found that clinicians navigate uncertainty using both standardised processes (e.g., triage) and flexible approaches (e.g., negotiation), depending on acuity and available resources (93). McKelvie’s theory similarly proposes that engaging with clinical uncertainty can foster greater professional risk tolerance, a view supported by studies showing that clinicians with higher risk tolerance are less likely to admit patients unnecessarily in emergency settings (19–21).

Finally, the extent to which clinical risk is effectively distributed within and across teams depends on organisational culture and interprofessional trust (IPT 3). Clearly defined roles, mutual respect, and collaborative working practices facilitate shared responsibility for clinical decision-making. In contrast, risk-averse organisational environments characterised by ambiguity or rigid hierarchies can erode confidence and constrain professional autonomy, increasing the likelihood of hospital admission as a default option. Evidence suggests that healthcare professionals in organisations with a higher risk appetite for out-of-hospital care feel more empowered to manage complex or high-acuity cases in the community (106). These organisations often support more flexible, collaborative, and person-centred approaches. In contrast, those with lower risk appetite tend to favour hospital admission, underpinned by stricter protocols, hierarchical governance, and lower thresholds for escalation.

**Figure 1.**
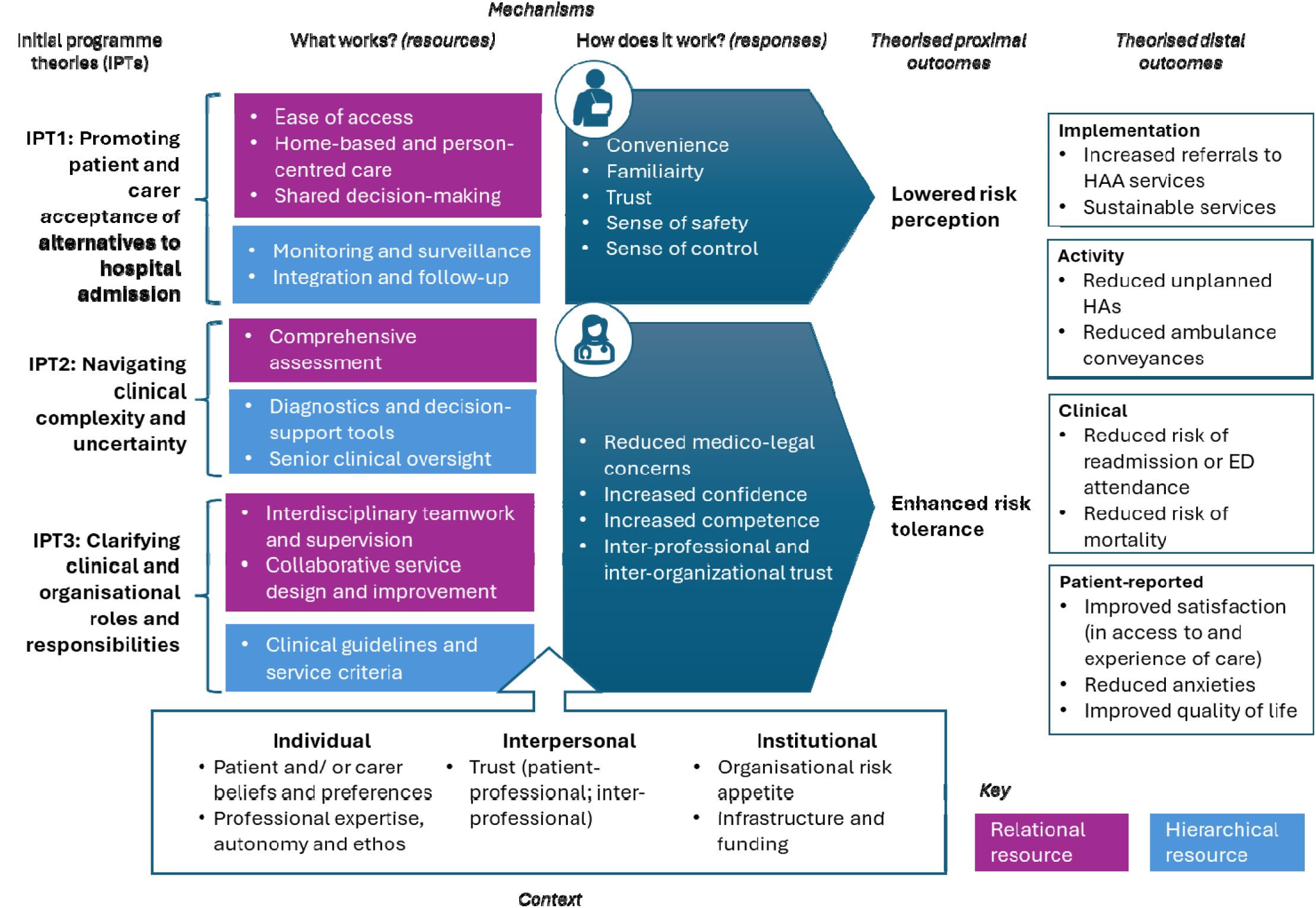
Summary Diagram of Initial Programme Theories of Hospital Admission Avoidance Interventions.

Across all IPTs, trusting relationships, between patients and providers, within teams, and across organisations, enabled individuals to act with greater certainty in uncertain situations. This finding aligns with social theories that position trust as a mechanism for reducing uncertainty (103). In healthcare settings, trust supports risk tolerance by empowering HCPs to make decisions in complex cases, while also facilitating inter-organisational collaboration and shared accountability (18, 104). It can also increase patients’ and carers’ confidence in non-hospital options by reinforcing the reliability and safety of these alternatives (105).

### Implications for policy and practice

This synthesis highlights several key considerations and inherent tensions in navigating and managing risk in relation to hospital admission avoidance. Firstly, there is a need to balance improving equitable access to HAAIs for people with UCSCs while simultaneously managing demand to preserve limited healthcare resources. Careful and thoughtful service design is required to ensure that HAAIs offer acceptable and equitable alternatives without inadvertently increasing demand or compromising the intended scope of services. Secondly, enabling healthcare professionals to confidently engage with clinical complexity and uncertainty in their decision-making, while maintaining person-centred care, requires sufficient time, appropriate clinical expertise, and an appropriate level of professional autonomy. Thirdly, although interdisciplinary teams must often collaborate to share clinical risk and deliver high-quality care, a clearly defined organisational risk appetite, supported by strong governance, is essential to create the conditions that empower clinicians to make consistent and confident decisions to avoid hospital admissions.

This synthesis demonstrates that effective HAAIs depend on balancing hierarchical strategies to manage risk with relational strategies to navigate risk. The optimal balance varies by context: high-acuity or time-sensitive cases may require stronger hierarchical oversight, while complex or preference-driven situations benefit from adaptable, relational approaches (93). Overreliance on either approach can undermine effectiveness, as supported by existing evidence on healthcare risk governance (106, 107). Optimal outcomes occur when hierarchical structures define standards of ‘what good looks like’ without constraining the relational practices necessary for delivering ‘good care’ (108). To support sustainable implementation, policy makers and service leaders must ensure that HAAI strategies explicitly address the risks experienced by patients, carers, and healthcare professionals across varied settings.

### Study limitations

This realist review draws on recent empirical literature and stakeholder input to provide a broad, current understanding of how HAAIs work for different groups. However, its non-systematic approach may have missed relevant information affecting interpretation. Consistent with realist methodology, studies were selected for causal relevance rather than methodological hierarchy. While findings were refined iteratively with stakeholders, causal links and directionality cannot be confirmed, and alternative explanations are possible. Results should be interpreted cautiously until empirically tested.

Future research will aim to test the resource-response interactions identified here, examine which contextual factors are most strongly associated with effective HAAIs and whether risk tolerance and risk perception are proximal to intended outcomes. For example, validated measures of risk and uncertainty tolerance could be used to quantify their relationship with HAAI outcomes across different local health system and service contexts(19, 110).

## Conclusion

This realist review advances understanding of how hospital admission avoidance interventions (HAAIs) work, for whom, and in what contexts. By synthesising empirical evidence, stakeholder insights, and middle-range theory, we developed three initial programme theories that explain how key mechanisms, particularly those related to navigating and managing risk, shape the effectiveness and sustainability of HAAIs. Our findings highlight that patients, carers, and healthcare professionals all navigate perceived or actual risk when engaging with HAAIs. Effective HAAIs rely on striking an appropriate balance between hierarchical strategies, such as protocols, oversight, and governance, and relational strategies that foster trust, shared understanding, and flexible, person-centred care. This balance must be context-specific and responsive to the acuity of clinical need, the complexity of care, and individual and organisational capacity for risk tolerance.

Embedding HAAIs successfully requires recognising the wider system within which they operate. Organisations delivering HAAIs must establish a clear risk appetite and ensure the appropriate infrastructure, operational settings, and resources are in place. Mutual trust between patients, carers and professionals and between professionals is essential and individual differences in attitudes towards HAA must be taken into account. Further empirical research is needed to test the proposed context-mechanism-outcome configurations and determine how risk-related mechanisms influence longer-term outcomes. Nonetheless, this synthesis offers a theoretically grounded foundation for designing and implementing HAAIs that are both effective and resilient in diverse health system contexts.

## Supporting information

Supplementary Material - Search Strategies and PRISMA

## Data Availability

The datasets used and/or analysed during the current study are available from the corresponding author on reasonable request.

## List of abbreviations

CGA: Comprehensive Geriatric Assessment
ED: emergency department
GP: General Practitioner
HA: hospital admission
HAA: hospital admission avoidance
HAAI: hospital admission avoidance interventions
HAH: Hospital at Home (service)
HCP: Health and Care Professionals
UCSC: urgent care sensitive conditions
UCR: Urgent Community Response (service)
VW: Virtual Ward (service)

## Declarations

### Ethics approval and consent to participate

Not applicable

### Consent for publication

Not applicable

### Authors’ contributions

MB and HB conceived the study. CM developed the protocol with input from MB, AH and HB. CM conducted the search, screening and synthesis, with regular discussions with MB. CM and MB carried out stakeholder engagement activities. AH addressed any discrepancies in study inclusion. CM wrote the manuscript and MB, AH and HB provided comments on drafts. All authors reviewed the final version of the manuscript.

## Acknowledgements

Justin Jagosh (JJ) was consulted during protocol development and during the early stages of synthesis.

