## Supplementary Material - Search Strategies and PRISMA for "Risk tolerance and risk perception as determinants of successful and sustainable hospital admission avoidance interventions: a realist review"

Supplementary Table 1. Search strategy for MEDLINE

| Set | Search Statement |
| --- | --- |
| 1. | (avoid* adj3 admission*).mp. |
| 2. | (appropriate adj3 admission*).mp. |
| 3. | (prevent* adj3 admission).mp. |
| 4. | (unnecessary adj3 admission*).mp. |
| 5. | (avoid* adj3 hospital*).mp. |
| 6. | (prevent* adj3 hospital*).mp. |
| 7. | Ambulatory Care Sensitive Conditions.mp. or exp Ambulatory Care Sensitive Conditions/ |
| 8. | (general practice or GP).mp. or exp General Practice/ |
| 9. | exp health facility planning/ |
| 10. | exp community health planning/ |
| 11. | (community adj2 intervention).mp. |
| 12. | ambulatory care.mp. or exp Ambulatory Care/ |
| 13. | *Community Health Services/ |
| 14. | Home Care Services/ |
| 15. | (home adj3 care).mp. |
| 16. | Telemedicine/ |
| 17. | (remote adj3 care).mp. |
| 18. | virtual ward.mp. |
| 19. | (hospital adj2 home).mp. |
| 20. | (frailty adj2 service).mp. |
| 21. | (frailty adj2 unit).mp. |
| 22. | (frailty adj2 ward).mp. |
| 23. | urgent community response.mp. |
| 24. | rapid response.mp. |
| 25. | (advice adj2 guidance).mp. |
| 26. | (walk-in adj2 cent*).mp. |
| 27. | (drop-in adj2 cent*).mp. |
| 28. | out-of-hours.mp. |
| 29. | expanded hours.mp. |
| 30. | (enhanced adj3 triage).mp. |
| 31. | single point of access.mp. |
| 32. | palliative care.mp. or Palliative Care/ |
| 33. | crisis intervention.mp. or Crisis Intervention/ |
| 34. | emergency mental health.mp. |
| 35. | (emergency adj2 social).mp. |
| 36. | reablement.mp. |
| 37. | protocol.m_titl. |
| 38. | (unnecessary adj3 hospital*).mp. |
| 39. | admitting department.mp. or Admitting Department, Hospital/ |
| 40. | emergency department.mp. or *Emergency Service, Hospital/ |
| 41. | 8 or 9 or 10 or 11 or 12 or 13 or 14 or 15 or 16 or 17 or 18 or 19 or 20 or 21 or 22 or 23 or 24 or 25 or 26 or 27 or 28 or 29 or 30 or 31 or 32 or 33 or 34 or 35 or 36 or 39 or 40 |
| 42. | (unnecessary adj3 visit*).mp. |
| 43. | (avoid* adj3 visit*).mp. |
| 44. | (prevent adj3 visit*).mp. |
| 45. | (appropriate adj3 visit*).mp. |
| 46. | 1 or 2 or 3 or 4 or 5 or 6 or 7 or 38 or 42 or 43 or 44 or 45 |
| 47. | 46 and 41 |
| 48. | 47 not 37 |
| 49. | limit 48 to (english language and yr="2019 -Current") |

Supplementary Figure 1. PRISMA study selection diagram

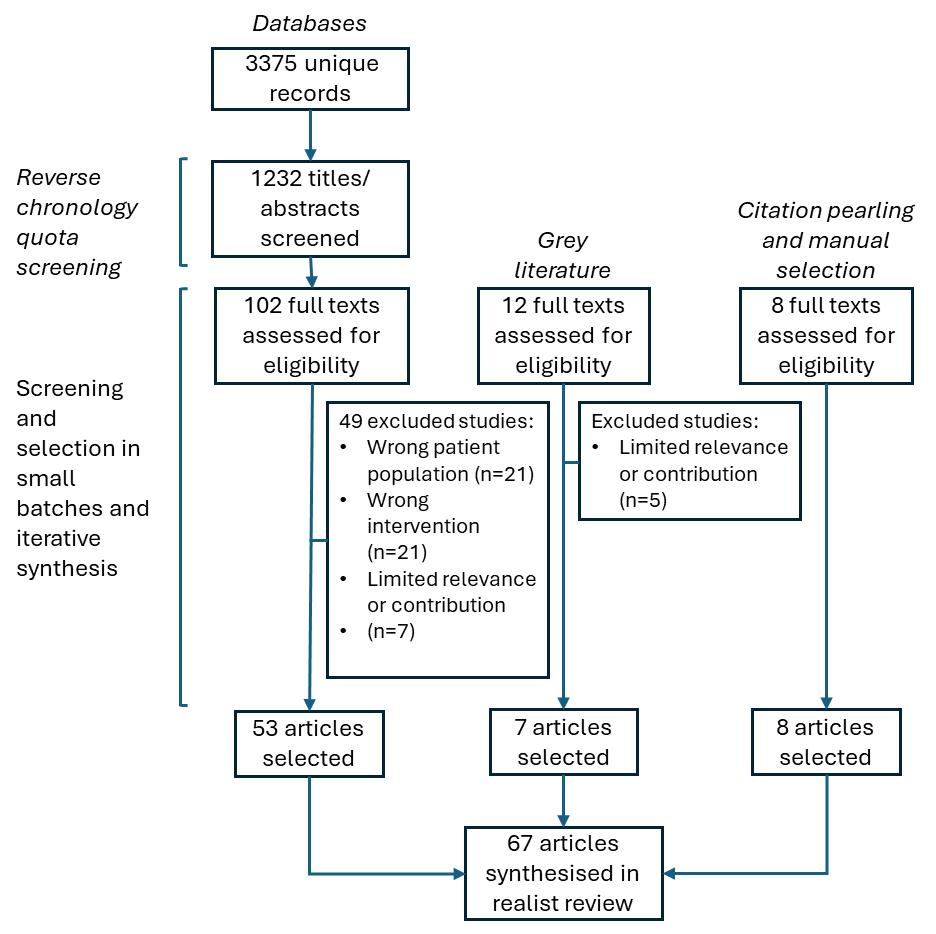
